# HBV prevalence in Sub-continental countries: a systematic review and meta-analysis

**DOI:** 10.1101/2023.04.23.23288908

**Authors:** Sam Hogan, Andrew Page, Sameer Dixit, Kate McBride

## Abstract

**Background:** Hepatitis B virus (HBV) is a major source of disease burden worldwide, especially in developing nations. Although vaccine programs exist to control infections, certain sub-populations around the world continue to have very high prevalence of HBV infection.

**Methods:** A systematic search of studies of HBV published after 2010 was conducted for India, Pakistan, Bangladesh, Nepal, Sri Lanka and Bhutan. Each paper was independently screened for risk of bias and inclusion. Data were extracted from included studies before being analysed to estimate pooled prevalence, and to conduct sub-group analyses. Random-effects models were used for estimating summary prevalence due to a high level of heterogeneity between studies, and funnel plots were combined with Egger’s test to assess publication bias. Meta-regression was conducted to investigate sources of between-study heterogeneity.

**Results:** The pooled prevalence of HBV across all studies was 4% (95% CI 0.02, 0.06). For countries with multiple studies, the pooled prevalence of HBV was 3% (95% CI 0.02, 0.05) for India, 6% (95% CI 0.04, 0.10) for Pakistan, 5% (95% CI of 0.02, 0.12) for Bangladesh, and 1% (95% CI 0.00, 0.08) for Nepal. There was some evidence of publication bias, and a high level of heterogeneity across studies. Risk of bias analysis found most studies to be of fair or moderate quality.

**Conclusions:** The prevalence of HBV among sub-continental countries was higher than the global average but was not as high as some other regions. Countries with greater numbers of displaced persons had higher prevalence of HBV, with a wide range of prevalence between subpopulations likely reflecting differential uptake, and implementation, of vaccination programs.

## INTRODUCTION

Hepatitis B is an infection caused by the hepatitis B virus (HBV) and can lead to severe complications in infected individuals. HBV infections are a major cause of health problems worldwide, with both chronic and acute infections presenting different symptoms and complications. In 2015, HBV was the cause of an around 887,000 deaths worldwide and there were an estimated 257 million people living with a chronic infection (1), although these numbers differ geographically (2). Chronic HBV infections can cause cirrhosis of the liver and in some cases, hepatocellular cancer. Acute HBV infections have no specific treatment, with the majority of treatments aiming to reduce symptoms and managing the discomfort of the infected individual. Symptoms of acute HBV infections can include dehydration, diarrhoea and vomiting. Children, especially newborns, are extremely vulnerable to developing chronic HBV infections, as the majority of those infected within the first year of life will go on to develop a chronic infection. Although the chance of developing chronic infection reduces with age, 30-50% of those infected with HBV before the age of six will develop a chronic infection (1). This is especially problematic, as one of the most common routes of transmission of HBV infections is vertical transmission from mother-to-child or direct transmission from an infected child to others via exposure to infected blood from the child (1, 2). Other methods of transmission include exposure to infected blood or other bodily fluids, sexual transmission and intravenous drug use via sharing needles or use of unsterilised needles (2, 3). Contaminated razors can also be a method of transmission, which is problematic in parts of the world where barbershops play important roles both socially and culturally (4).

Although a vaccine is available for HBV that provides protection from infection, high levels of vaccination coverage are not ubiquitous in many parts of the world. The rate of HBV infection among general populations also varies from country to country, as vaccination regimes and protocols differ between settings. Different regions around the world have varying prevalence of HBV, with the World Health Organization (WHO) Western Pacific region and African region reporting the highest prevalence (1). Many developing countries began implementing HBV vaccination programs relatively recently, which have shown some success in reducing the prevalence of HBV in younger populations (5, 6).

In this systematic review and meta-analysis, we sought to ascertain the prevalence of HBV infection among populations within subcontinental countries. For studies which included HBV vaccination status as a variable, the vaccination rates were also examined as a secondary aim. Additionally, subgroups were assessed to determine which factors increase risk of HBV infection and reduce the chance of being vaccinated for HBV. Pre- and post-vaccination cohorts were also examined to assess differences in HBV prevalence that may be attributable to vaccination programs within settings where data was available.

## METHODS

### Study Settings

The countries chosen for inclusion in this study were those of the Indian Subcontinent, namely India, Pakistan, Bangladesh, Nepal, Sri Lanka and Bhutan. The Maldives were also included in the search strategy, however no relevant papers were located. The Indian Subcontinent was chosen as the region of focus for this review as the countries are all densely populated low and middle income countries, share a close national history, and there is relatively free movement across borders for many of these countries. Additionally, as vaccination programs have been introduced in the selected countries from the early 2000s it is important to examine changes in prevalence which may be occurring, especially as there have been different prevalence levels reported from certain subgroups which are higher than those of national averages and other international contexts (7, 8). Thus, examining vaccination coverage within the populations of interest was a secondary aim of this review. General populations from both rural and urban populations were also chosen for inclusion in this systematic review, as these settings have different risk factors that may increase or decrease the chance of HBV infection.

### Study Design and Protocol Registration

The protocol of this systematic review and meta-analysis was designed based on the Preferred Reporting Items for Systematic Reviews and Meta-Analysis Protocols (PRISMA-P) Guidelines (9). The protocol for this systematic review was registered in Prospero prior to the initial search (protocol registration number of CRD42020215743).

### Search Strategy and Inclusion criteria

A systematic search was conducted using multiple databases; PubMed, Embase, Medline and the Cochrane Library. Web of Science was also used, but all articles returned from this search were duplicated in the results from the other databases. The search strategy restricted results to only those published after 2010, to ensure recency and increase the relevance of the papers. Some of the countries of interest had previously had systematic reviews focusing on HBV which included results up to 2010, so this rationale was also considered when defining this restriction.

The search strategy used a combination of keywords, search symbols and MeSH terms to strengthen the search. The full list of search terms and how they were combined is included in Appendix X. The search was conducted by the first author (SH), and the results from each database combined. Following this, the titles were scanned to assess relevance, which was done by assessing whether the terms “HBV” or “prevalence” were included in the titles. Those titles which did not include one of these two terms were excluded from the next phase, where abstracts were independently assessed by each reviewer (SH, AP and KM) for relevance. When assessing risk of bias, full text reviews were conducted on those papers chosen from inclusion following the abstract scan.

### Inclusion and Exclusion Criteria

Inclusion criteria were studies conducted in countries in the sub-continent, published after 2010, among participants with no underlying health conditions and of any age, and reported HBV prevalence with a valid testing method. Studies were restricted to English language articles, and with full text available. n. Another exclusion criterion was that participants could have no other underlying health issues (e.g., thalassemia patients, hepatocarcinoma patients) as these populations would likely not be representative of the general populations of identified countries.. All studies were required to be a prevalence study with a validated method of HBV diagnosis, ranging from rapid test kits to lab confirmation. No specific study design was specified, however due to the nature of the outcome of interest most studies were cross-sectional study designs. A secondary aim of the study was also to assess vaccination coverage within the study populations, however this was not included in many of the papers focussing on prevalence.

Almost all included studies used the same, or very similar, methods for identifying HBV prevalence within the respective study populations. The only other inclusion criteria was that the full text was available. The search strategy is as listed in Supplementary Table 1 (formatted for PubMed). Supplementary Figure 1 shows the flow chart of selecting the papers for inclusion in this review, based on the PRISMA Guidelines.

### Data Extraction

Data extracted from each study included study design, laboratory methods, age, sex, geographic location, literacy level, and vaccination status (where available), to allow for subgroup analysis by these variables.

### Quality Assessment

Following selection of studies that met the inclusion criteria papers, data extraction was performed for all relevant variables. The Joanna Briggs Checklist for Prevalence Studies (10) was used to assess the quality of the studies and risk of bias. This tool was chosen as it has been shown to be a valid method of assessing prevalence studies, while other tools such as the Cochrane Risk of Bias tool are less appropriate for this type of study design. All studies were assessed for risk of bias by one author (SH), and two other authors (AP and KM) assessed 50% of the papers each. Once risk of bias had been assessed, data analysis was performed on the selected papers. There was some disagreement between reviewers when classifying the quality of the studies (disagreement on 24/57 papers, 42%), however these issues were resolved by the 3^rd^ independent reviewer (KM or AP).

### Data synthesis and analysis

For pooled prevalence, random effects models were used as it was likely that variance between identified studies was due to more than only selection bias or sampling errors. Heterogeneity between studies was assessed using the I^2^ statistic. 95% confidence intervals were also given for the meta-analyses, with weights of each of the studies also shown. Publication bias and small-study effects were assessed using, funnel plots and the Egger’s test statistic. Subgroup analysis and meta-regression were used to assess sources of between study heterogeneity between studies. Meta-regression assessing the impact of a number of variables on the likelihood of being infected with HBV were used to identify common risk factors within the datasets. Meta-regressions were performed using country, sex, location, study quality, and year of publication, and source population of the studies. Literacy level was also used for a smaller meta-regression using those papers in which it was a recorded variable. Subgroup analysis was possible using sex, country, geographic location (i.e., rural or urban), age groups, study quality, and literacy levels. These categories were also used for meta-regressions when possible. Age group definitions varied across the included studies, therefore subgroups were created around common age-group definitions across studies. Similarly, literacy and illiteracy were clearly defined in some studies (7, 11, 12) but not in others. Biological sex was the only variable which was consistently present in each of the studies, however as some studies focussed exclusively on either males or females, not all studies were included in the analysis for this subgroup analysis.

Data analyses were conducted using R (13) and RStudio (14) using the “metafor” package (15), while some figures were visualised from the same dataset using the Stata software package.

## RESULTS

### Prevalence of HBV

Of the 56 papers that met the inclusion criteria, 23 were based in India, 19 in Pakistan, 8 in Bangladesh, 4 in Nepal and 1 each from Sri Lanka and Bhutan. The pooled prevalence of HBV for all included studies was 4% (95%CI 0.02, 0.05) (Table 1, Supplementary Figure 2). The prevalence of HBV for Sri Lanka and Bhutan was <1% (95%CI 0.00, 0.08) and 1% (95%CI 0.01, 0.02) respectively, and based on single studies. The pooled prevalence of HBV was 3% (95%CI 0.02, 0.05) in India, 6% (95%CI 0.04, 0.10) in Pakistan, 5% (95%CI 0.02, 0.12) in Bangladesh, and 1% (95%CI 0.00, 0.08) in Nepal. The majority of the included studies were cross-sectional studies, with the exception of one prospective study, 2 retrospective studies and 8 studies with poorly described study design.

**Table 1.**
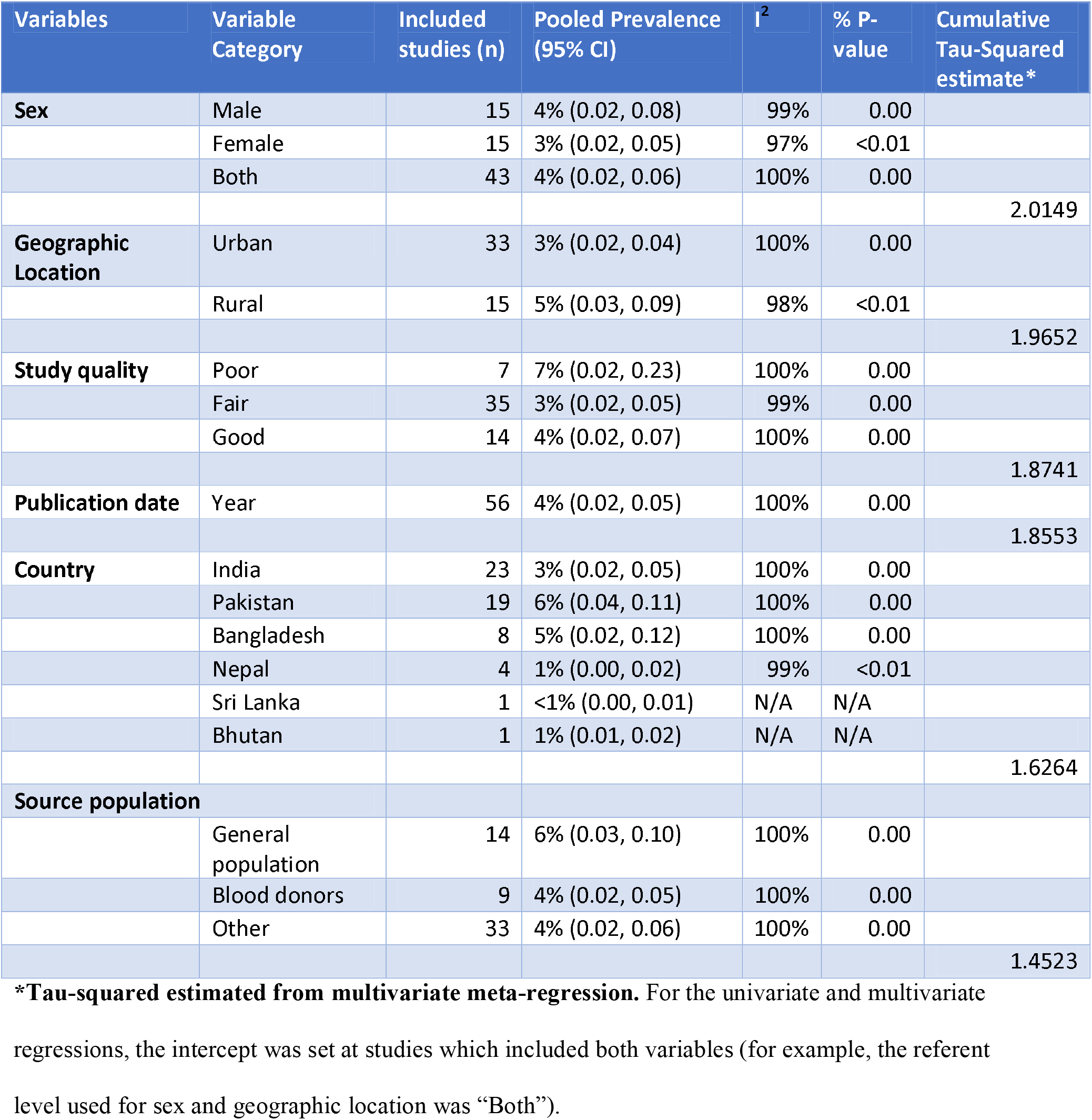
Subgroup analysis assessing pooled prevalence of HBV and sources of heterogeneity.

There was also marked within- and between-country differences in HBV prevalence. For example, the highest prevalence for India was recorded by Kuriakose and Ittyachen (16) who were investigating a rural area of Kerala. Although the sample population was relatively small, the HBV prevalence was 30% (95% CI = 0.24, 0.37), far higher than the pooled prevalence of 3% (95% CI = 0.02, 0.05). The total population of all included studies was 715,482 individuals from across the 6 countries, with 16,994 positive results for HBV. Heterogeneity between the studies was very high, with *I*^2^ = 100% and τ^2^ = 2.1154 for the pooled prevalence of all studies, and was also very high for country-specific pooled prevalence. This high level of heterogeneity is likely due to the highly varied population sizes and locations of the studies, as many studies were conducted using a similar design.

### Vaccination coverage

Vaccination coverage was assessed in several of the studies, however vaccination coverage was predominantly ascertained via participant self-report. The majority of those studies that assessed vaccination coverage found the completion rate for the full 3 dose course was less than 90%, with the pooled average being 58% across these studies (5, 6, 17-22). There were also some studies which included data from study populations born before national vaccination programs were initiated compared to those born after the programs were introduced (6). For example, a study based in Bhutan found that the prevalence of HBV was far lower in the population who had been born after the vaccination program had been implemented (6). Additionally, in a Nepal-based study, the prevalence of HBV was markedly lower in the post-vaccination cohort of children when compared to a pre-vaccination cohort (5).

### Subgroup Analysis

The pooled prevalence of HBV among adult males was 4% (95%CI 0.02, 0.08), established using the random effects model. For adult females, including pregnant women, the pooled prevalence of HBV was 3% (95%CI 0.02, 0.05). The pooled prevalence of HBV among the rural only populations was 5% (95%CI 0.03, 0.09), while urban populations had a lower prevalence of 3% (95%CI 0.02, 0.05). When comparing literate vs illiterate populations, the pooled prevalence of HBV was 9% (95%CI 0.03, 0.23) compared to 18% (95%CI 0.13, 0.23), although the number of articles which recorded this variable was small (n = 3). The results of the subgroup analyses are shown in Table X? below. Some subgroups examined had a markedly higher prevalence of HBV than the national averages for the general population, although some subpopulations reflected the expected prevalence for the countries in which they were located. For example, in a study which examined individuals in Northern Pakistan which had been displaced by conflict (7) the prevalence of HBV was far higher (21% compared to the pooled prevalence of 6%).

τ^2^ was used to determine variation of the true effect within subgroups examined in the meta-analysis. Meta-regressions suggested that the main source of between study variation was country of study setting (Tau-Sq = 1.6264), followed by the source population (Tau-Sq = 1.4523), and study quality (Tau-Sq = 1.8741) (Table 1). These variables were associated with the biggest change in Tau-Sq within the multivariate model. The variables selected within the multivariate regression also accounted for the majority of between study variation.

### Risk of Bias

The majority of the studies included in this review were of fair quality (n = 35), with only a few rated as poor (n = 7). Fourteen of the papers were rated as being of good quality, as these papers reported on all key aspects of their methodology. A summary of the study characteristics of each of the included papers can be found in Supplementary Table 2 in the appendix. There was some evidence of publication bias, as indicated by funnel plots for all subgroups (Supplementary Figure 3 in the Appendix), with Hedge’s G values indicating significant asymmetry.

## DISCUSSION

The Indian subcontinent is a region with a low overall prevalence of HBV infection, however between, and within, countries there remains wide variation in HBV prevalence. Understanding this regional variation is important to understand the performance of immunisation programs and potential priorities for health system strengthening within this geographic region. Nepal, for example, began widespread distribution of the HBV vaccine in 2002, although the prevalence of HBV within the country was already relatively low, with prevalence estimated to be between 2-4% (5). In comparison, other countries within the subcontinent have a higher prevalence, with India (4-7% prevalence) being the highest (23). Pakistan has recorded a prevalence of 3-5% in their general population (11), while Bangladesh has an estimated prevalence of 5.4% (24). Bhutan and Sri Lanka both have lower prevalences, with an estimated prevalence of approximately 2% in each country (6, 25). These lower prevalence levels may be due to various geographic and cultural features. Sri Lanka shares no land borders with other countries, while Bhutan is relatively isolated due to the dense forest and mountainous terrain which makes up the majority of the country.

Various subgroups within the general population of each of these countries have been shown to have a higher prevalence of HBV based on a variety of risk-taking behaviours, for example use of intravenous drugs. Although some risk-taking behaviours are clear, others are related to lifestyle or living conditions which are outside of the control of the individual, such as being displaced by internal conflicts (7, 26).

The prevalence of HBV among the populations in the papers included in this systematic review varied significantly. While some of the study populations reflected the expected prevalence for the country in which the study was situated, other subpopulations from the same country had far higher prevalence of HBV. While some of these variations were due to clear at-risk behaviours, other variations had less clear aetiology. Additionally, vaccination programs appear to have had variable success across these settings, with some reporting coverage far lower than the targeted proportion. However, studies did show that vaccination programs have been successfully introduced in some of the settings. In these studies, there was a clear reduction in the prevalence of HBV in pre- and post-vaccination program groups. HBV prevalence in general is lower at present than in the past, however there are still many at-risk groups with far higher prevalence than the general population. Usually, these populations are disadvantaged in some way, for example being displaced by war (26).

In general, studies set in urban locations had a lower HBV prevalence than those in more rural areas. This could be due to a number of factors such as decreased access to healthcare, differences in education levels and higher frequency of other at-risk behaviours. Study participants who were illiterate were also far more likely to have an HBV infection, which may be tied with overall education levels and health literacy, as they may be unaware of common behaviours which act as risk factors for HBV.

When examining which population groups studies focussed on, those focussing on the general population showed a higher pooled prevalence than any other group. Studies were classified as examining the “general population” if they did not restrict the sample population or did not focus on a specific feature of a population when sampling. Some populations were suspected of being at higher risk of HBV infection due to a variety of factors, but were still representative of the general population. However, the studies which examined “Other” populations focussed on populations which would not be representative of the wider population as a whole, such as medical students (19), healthcare workers (27) or the general population of a specific group of remote islands (20, 28). This meant that the variation between prevalence of HBV among each group varied greatly, as some reasons that these populations were not representative of the wider population placed them at greater risk of becoming infected, while others seemingly had a protective effect. The pooled prevalence of the “general population” group was 6% using the random effects model, while the “other” population subgroup had a pooled prevalence of 4%. The sample population with the lowest pooled prevalence of HBV were blood donors, which was expected.

Studies classified as “Fair” or “Good” after risk of bias assessment had similar prevalence levels of 3% and 4% respectively. However, studies classified as “Poor” had a far higher prevalence of 7%. This may be due to selection of study populations not representative of the general population (e.g. hospital patients with known hepatitis). However, the settings of some of these studies were likely to have impacted the quality of the studies themselves and may have made data collection more difficult, for example the studies focussing on rural areas or populations who had been displaced from their normal area of residence (7). It is unlikely that the difference is due to measurement error as almost all the studies utilised an enzyme-linked immunosorbent assay (ELISA) test to confirm HBV infection status, with some also utilising polymerase chain reaction (PCR) tests to additionally confirm diagnoses. Sampling issues may have affected several of the studies, as many were based on convenience samples. While follow up is not normally relevant in cross-sectional studies, sample selection should still be adequately described in all papers. The main issue with many of the papers was how representative the study population was of the wider population the studies were set in. Additionally, some papers had poor reporting on how study populations were recruited, what the response rates were and how sampling was performed.

It is likely that there is some level of publication or sample bias, as the high level of asymmetry shown in each of the funnel plots suggest that the prevalence from many of the studies fell outside the expected range. However, while some of these studies were potentially published for showing highly statistically significant results, this does not necessarily mean that all of the studies were. For example, the studies which were based on at-risk or vulnerable populations possibly showed higher prevalence simply because these populations are at an increased risk of HBV infection. Additionally, some of the studies with larger sample sizes showed a lower prevalence than that of the general population. However, some of the studies with the largest population sizes specifically examined blood donations. This population is likely to be healthier than the general population, so this may have affected observed results.

Other limitations common among the included studies were difficulties in recruiting a representative sample. Many of the studies focussed on specific subgroups rather than the general population, limiting generalisablity beyond those subgroups. While these subgroups are often at an increased risk of HBV, this is typically due factors not common to the general population (for example, refugee or intravenous drug user samples). However, some of the studies included in this review had substantial sample sizes, with smaller studies contributing a lower weight in pooled estimates.

## CONCLUSION

The results of this systematic review and meta-analysis of Hepatitis B virus infections within the countries of the Indian Subcontinent show that although the overall prevalence of HBV is decreasing in many countries, certain sub-populations remain at an increased risk of HBV infection. These groups in particular are those who have been displaced from their homes, as well as those who live in rural areas. Men also had a slightly higher risk of HBV infection than women, possibly a reflection of the different behaviours between sexes within country settings. Although there are some clear risk factors that increase the likelihood of HBV infection, the overall aetiology remains complicated, and vaccination programs remain critically important to help reduce the prevalence further.

## Data Availability

All data produced in the present work are contained in the manuscript.

## APPENDIX

### SUPPLEMENTARY TABLES AND FIGURES

**Supplementary Table 1.** Summary of the terms used for the systematic search of electronic databases.

|  | Search Terms |
| --- | --- |
| **Population:** People living countries within the Indian subcontinent | Subcontinent OR India OR Pakistan OR Nepal OR Bangladesh OR Sri Lanka OR Bhutan NOT China<br>**MeSH terms:** N/A |
| **Intervention:** Vaccination or no intervention | Vaccination* OR vaccin* OR no intervention OR immuni?ation<br>**MeSH terms:** Vaccination |
| **Comparison:** Rest of the world HBV prevalence | Prevalence OR trend AND Worldwide OR global OR world<br>**MeSH terms:** Prevalence |
| **Outcome:** Prevalence of HBV | HBV OR Hepatitis B* OR chronic HBV OR Hepatitis B infection OR HBV infection<br>**MeSH terms:** Hepatitis B virus, Hepatitis B, Hepatitis B Epidemiology |

**Supplementary Table 2.**
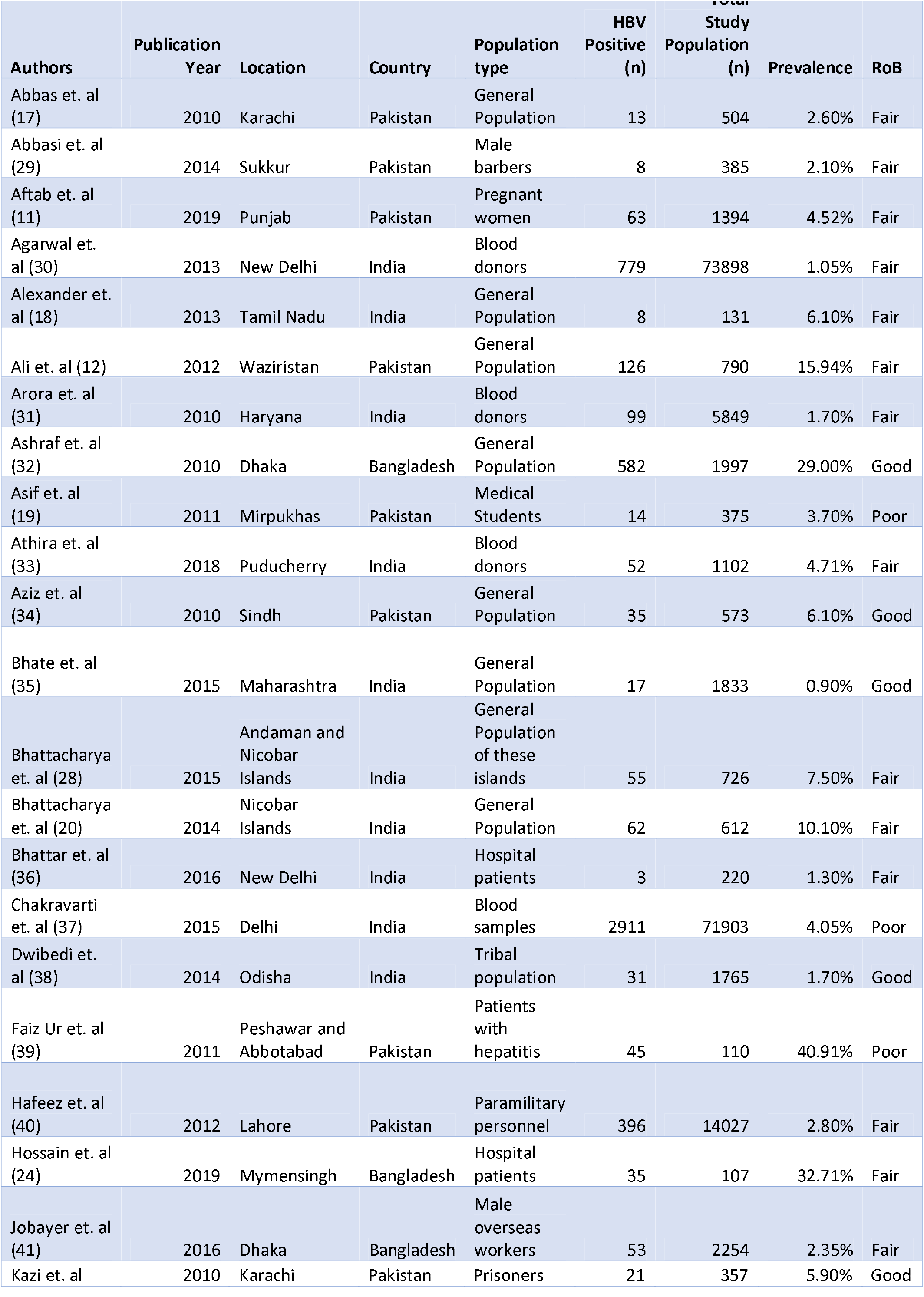
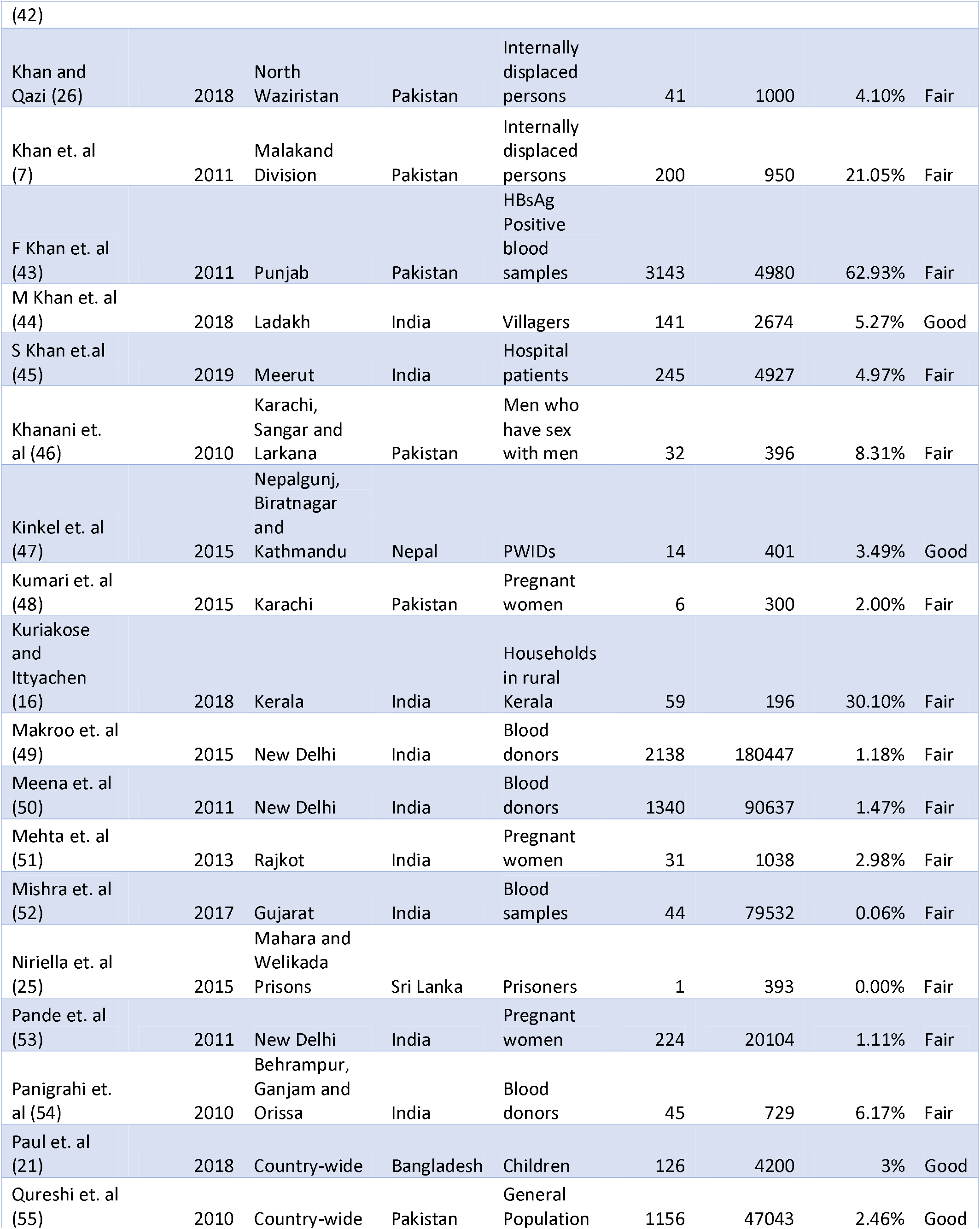
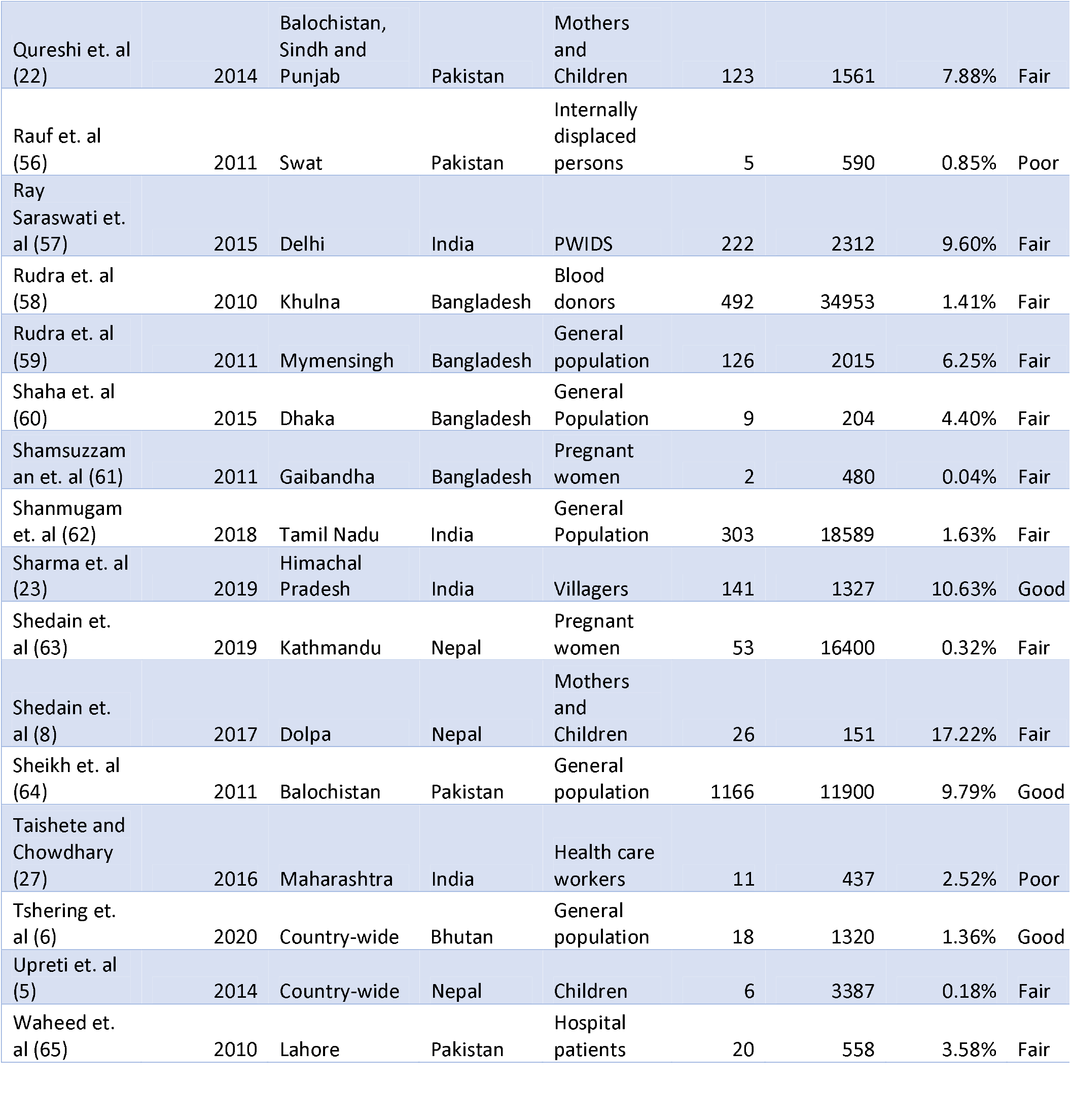
Summary Table of the studies included in this review. The variables listed are First author, year of publication, location, country of setting, population type, HBV positive, total population, HBV prevalence, and Risk of Bias assessment

**Supplementary Figure 1.**
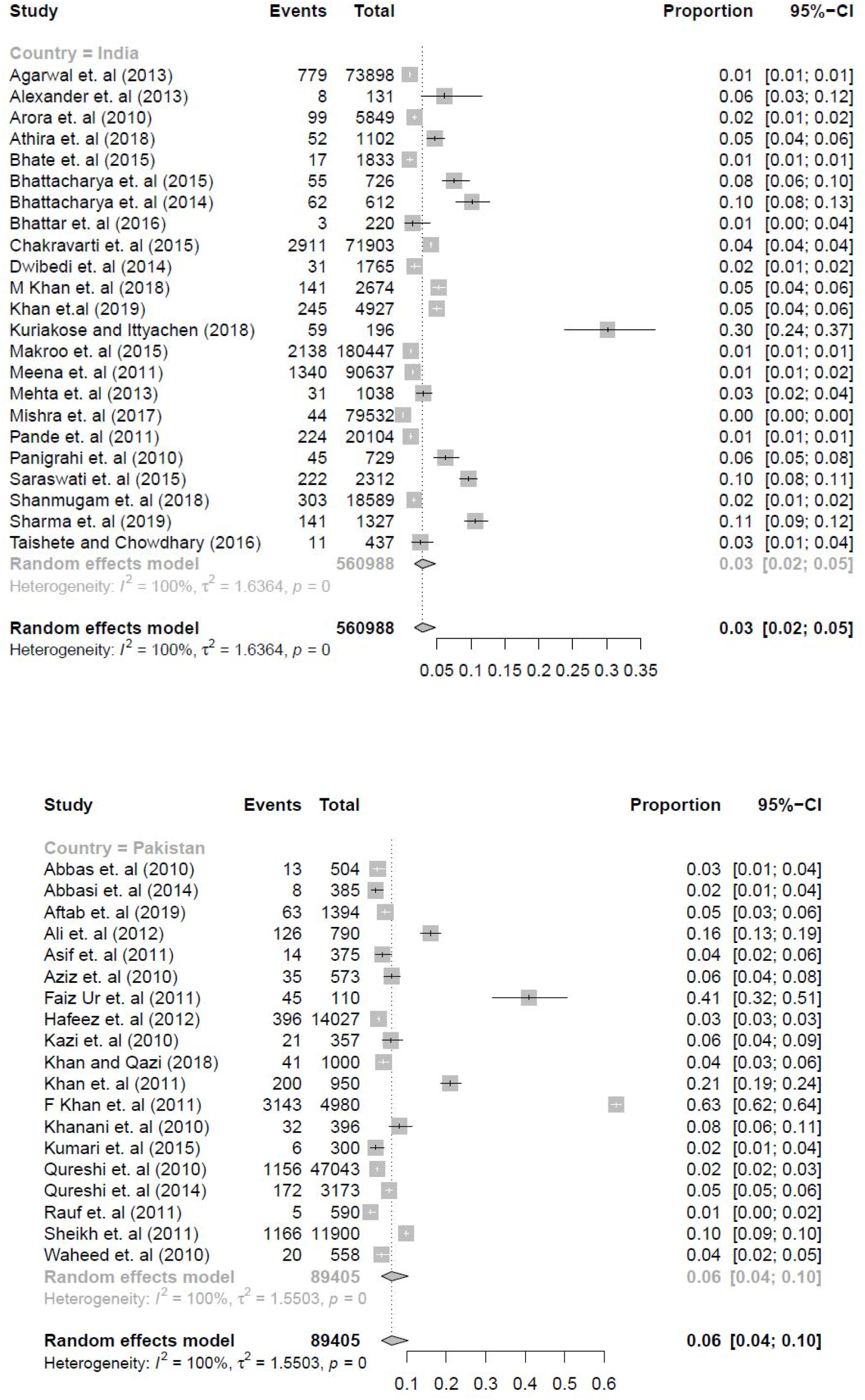
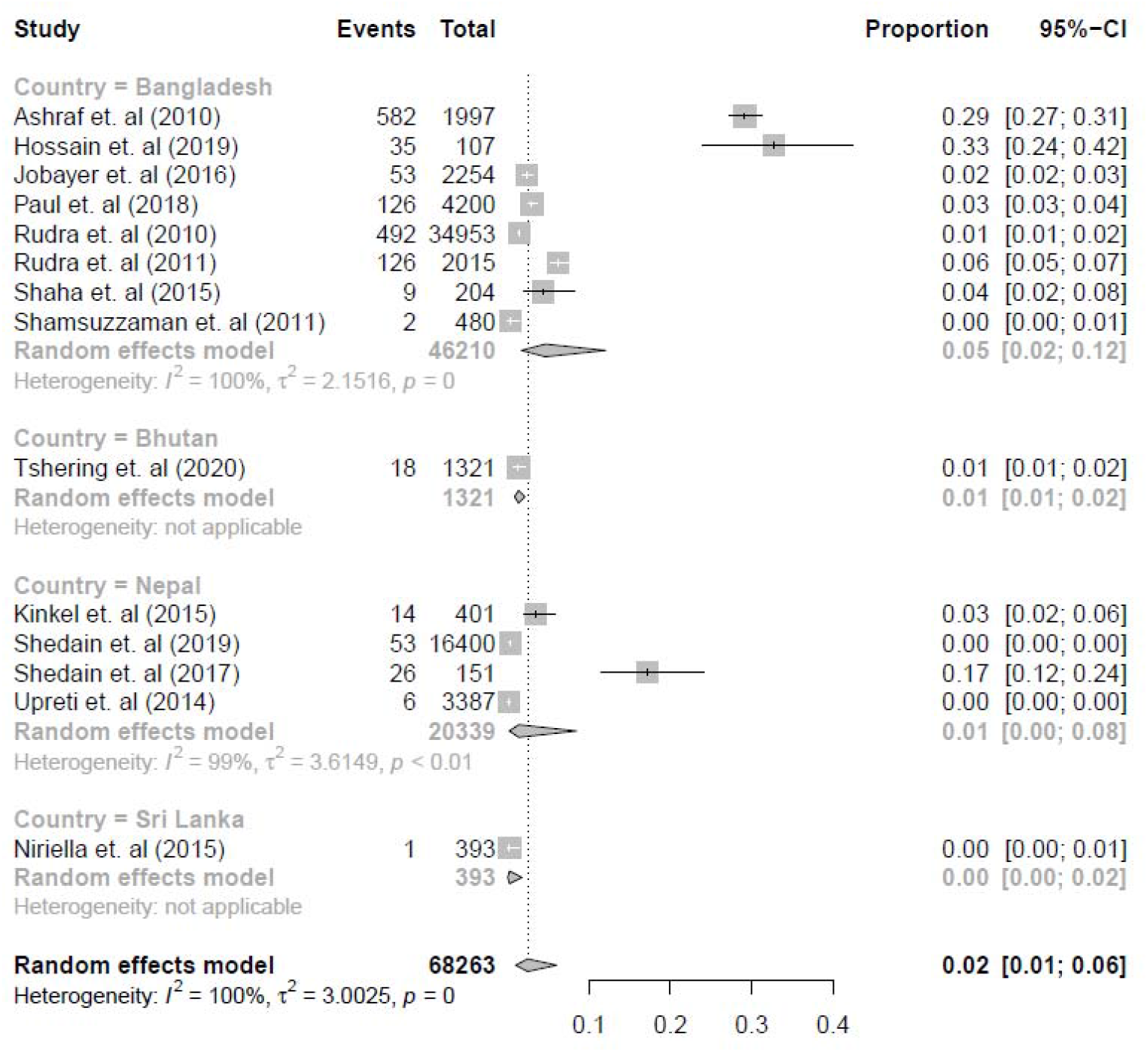
Forest plot of pooled prevalence stratified by country. The prevalence from each study is shown, as are the weights for the final overall pooled prevalence.

**Supplementary Figure 2.**
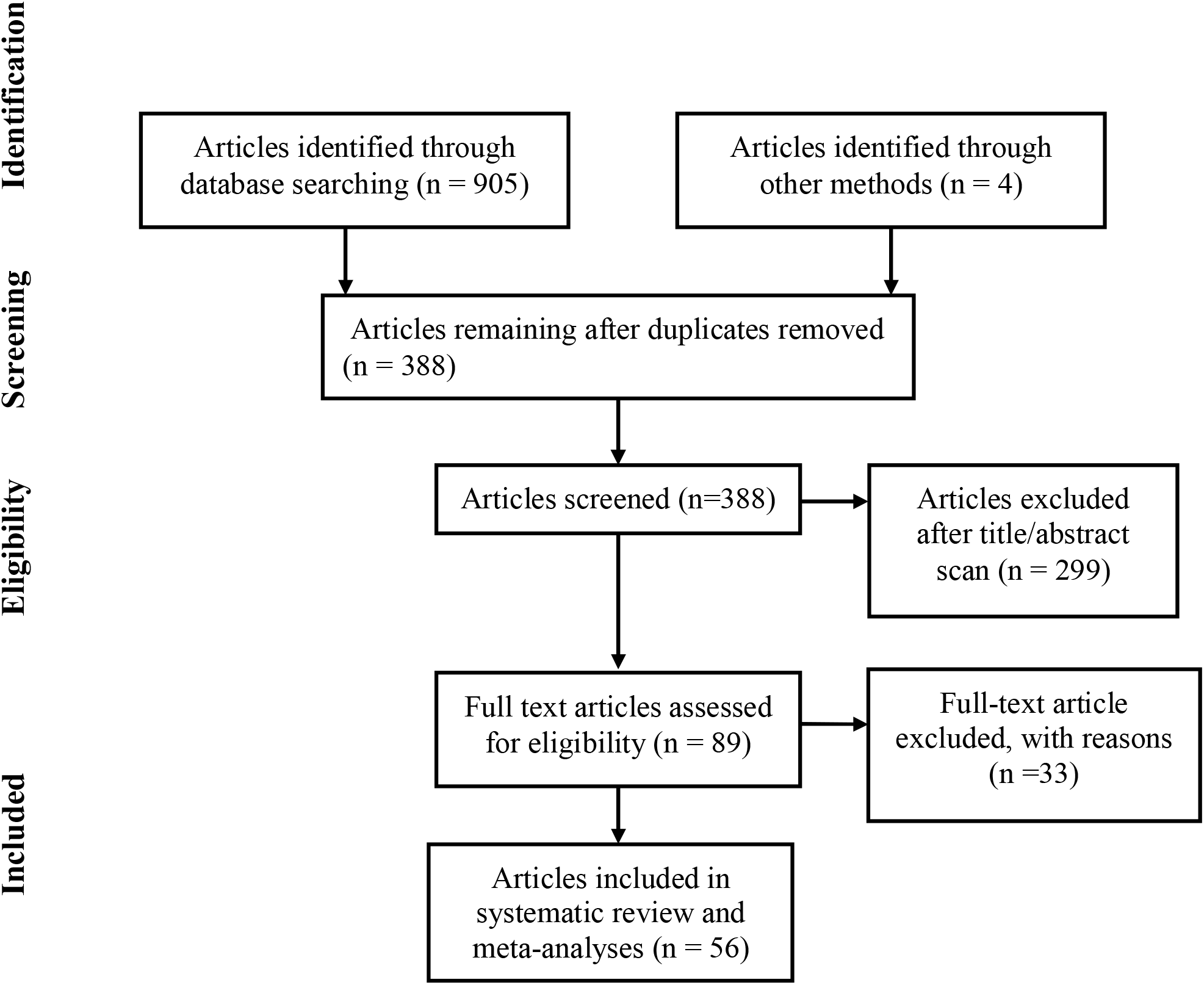
PRISMA Flowchart showing the selection process of the studies.

**Supplementary Figure 3.**
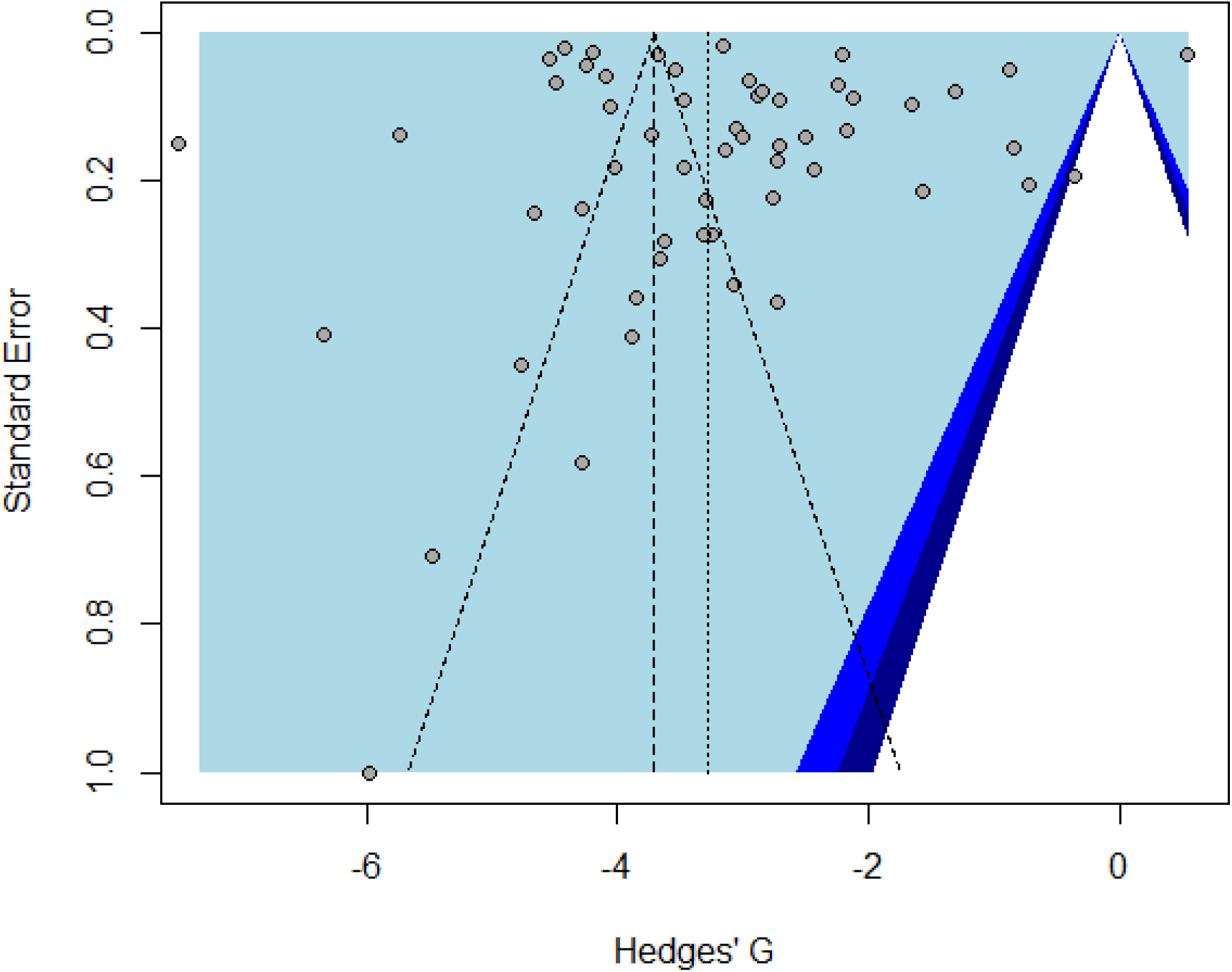
Funnel plot showing the chance of publication bias having an effect on study results. The shape of this plot indicates the large amount of variation between studies included in this review, and indicates the possibility that some publication bias exists.

## References

1. WHO. Hepatitis B WHO website: World Health Organisation; 2019 [Available from: https://www.who.int/news-room/fact-sheets/detail/hepatitis-b.

2. MacLachlan JH, Cowie BC. Hepatitis B virus epidemiology. Cold Spring Harb Perspect Med. 2015;5(5):a021410–a.

3. Degenhardt L, Peacock A, Colledge S, Leung J, Grebely J, Vickerman P, et al. Global prevalence of injecting drug use and sociodemographic characteristics and prevalence of HIV, HBV, and HCV in people who inject drugs: a multistage systematic review. The Lancet Global health. 2017;5(12):e1192–e207.

4. Shahid A, Nasim S, Memon AA. Insight and educational intervention concerning hepatitis among roadside barbers and their clients in Karachi, Pakistan. Journal of Infection in Developing Countries. 2013;7(2):125–9.

5. Upreti SR, Gurung S, Patel M, Dixit SM, Krause LK, Shakya G, et al. Prevalence of chronic hepatitis B virus infection before and after implementation of a hepatitis B vaccination program among children in Nepal. Vaccine. 2014;32(34):4304–9.

6. Tshering N, Dhakal GP, Wangchuk U, Wangdi S, Khandu L, Pelden S, et al. Prevalence of HBV and HCV infections, Bhutan, 2017: Progress and next steps. BMC Infect Dis. 2020;20(1):485.

7. Khan F, Akbar H, Idrees M, Khan H, Shahzad K, Kayani MA. The prevalence of HBV infection in the cohort of IDPs of war against terrorism in Malakand Division of Northern Pakistan. BMC Infect Dis. 2011;11:176.

8. Shedain PR, Devkota MD, Banjara MR, Ling H, Dhital S. Prevalence and risk factors of hepatitis B infection among mothers and children with hepatitis B infected mother in upper Dolpa, Nepal. BMC Infect Dis. 2017;17(1):667-.

9. Moher D, Liberati A, Tetzlaff J, Altman DG, Group P. Preferred reporting items for systematic reviews and meta-analyses: the PRISMA statement. PLoS medicine. 2009;6(7):e1000097–e.

10. Institute JB. The Joanna Briggs Institute Critical Appraisal tools for use in JBI Systematic Reviews; Checklist for Prevalence Studies. http://joannabriggs.org/research/critical-appraisal-tools.html: Joanna Briggs Institute; 2017.

11. Aftab M, Naz S, Aftab B, Ali A, Rafique S, Fatima Z, et al. Characterization of Hepatitis Delta Virus Among Pregnant Women of Pakistan. Viral immunology. 2019;32(8):335–40.

12. Ali A, Nisar M, Idrees M, Ahmad H, Hussain A, Rafique S, et al. Prevalence of HBV infection in suspected population of conflict-affected area of war against terrorism in North Waziristan FATA Pakistan. Infection, genetics and evolution : journal of molecular epidemiology and evolutionary genetics in infectious diseases. 2012;12(8):1865–9.

13. R Core Team. R: A language and environment for statistical computing.. R Foundation for Statistical Computing, Vienna, Austria; 2021.

14. RStudio Team. RStudio: Integrated Development for R. RStudio, Inc., Boston, MA; 2021.

15. Viechtbauer W. Conducting Meta-Analyses in R with the metafor Package. Journal of Statistical Software. 2010;36(3):1–48.

16. Kuriakose M, Ittyachen AM. An Investigation into the High Prevalence of Hepatitis B in a Rural Area of Kerala State, India: Hypothesis on Chrysops sp. (Diptera: Tabanidae) Transmission. BioMed research international. 2018;2018:4612472.

17. Abbas M, Hussain MF, Raza S, Shazi L. Frequency and awareness of hepatitis B and C in visitors of Hepatitis Awareness Mela. J Pak Med Assoc. 2010;60(12):1069–71.

18. Alexander AM, Prasad JH, Abraham P, Fletcher J, Muliyil J, Balraj V. Evaluation of a programme for prevention of vertical transmission of hepatitis B in a rural block in southern India. The Indian journal of medical research. 2013;137(2):356–62.

19. Asif M, Raja W, Gorar ZA. Hepatitis B vaccination coverage in medical students at a medical college of Mirpurkhas. JPMA The Journal of the Pakistan Medical Association. 2011;61(7):680–2.

20. Bhattacharya H, Bhattacharya D, Roy S, Sugunan AP. Occult hepatitis B infection among individuals belonging to the aboriginal Nicobarese tribe of India. Journal of infection in developing countries. 2014;8(12):1630–5.

21. Paul RC, Rahman M, Wiesen E, Patel M, Banik KC, Sharif AR, et al. Hepatitis B Surface Antigen Seroprevalence among Prevaccine and Vaccine Era Children in Bangladesh. The American journal of tropical medicine and hygiene. 2018;99(3):764–71.

22. Qureshi H. The evidence of mother to child transmission of hepatitis b virus infection in Pakistan and the need for hepatitis B immunization policy change. Journal of the Pakistan Medical Association. 2014;64(8):978.

23. Sharma RK, Shukla MK, Minhas N, Barde PV. Seroprevalence and risk factors of hepatitis B virus infection in tribal population of Himalayan district Lahaul and Spiti, India. Pathogens and global health. 2019;113(6):263–7.

24. Hossain MS, Alam MR, Hasan MI, Sharif JU, Kabir MA, Islam MA, et al. Prevalence of Serological Markers of Viruses in Patients of Acute Hepatitis. Mymensingh medical journal : MMJ. 2019;28(2):278–85.

25. Niriella MA, Hapangama A, Luke HPDP, Pathmeswaran A, Kuruppuarachchi KALA, de Silva HJ. Prevalence of hepatitis B and hepatitis C infections and their relationship to injectable drug use in a cohort of Sri Lankan prison inmates. The Ceylon medical journal. 2015;60(1):18–20.

26. Khan A, Qazi J. Risk factors and molecular epidemiology of HBV and HCV in internally displaced persons (IDPs) of North Waziristan Agency, Pakistan. JPMA The Journal of the Pakistan Medical Association. 2018;68(2):165–9.

27. Taishete S, Chowdhary A. Seroepidemiological survey of health care workers in Maharashtra. Indian Journal of Medical Microbiology. 2016;34(2):237–40.

28. Bhattacharya HB, Debdutta; Ghosal S. R.; Roy, Subarna; Sugunan, A. P. Status of hepatitis B infection - a decade after hepatitis B vaccination of susceptible Nicobarese, an indigenous tribe of Andaman & Nicobar (A&N) islands with high hepatitis B endemicity. The Indian journal of medical research. 2015;141(5):653–61.

29. Abbasi IN, Fatmi Z, Kadir MM, Sathiakumar N. Prevalence of hepatitis B virus infection among barbers and their knowledge, attitude and practices in the district of Sukkur, Sindh. International journal of occupational medicine and environmental health. 2014;27(5):757–65.

30. Agarwal N, Chatterjee K, Coshic P, Borgohain M. Nucleic acid testing for blood banks: an experience from a tertiary care centre in New Delhi, India. Transfusion and apheresis science : official journal of the World Apheresis Association : official journal of the European Society for Haemapheresis. 2013;49(3):482–4.

31. Arora D, Arora B, Khetarpal A. Seroprevalence of HIV, HBV, HCV and syphilis in blood donors in Southern Haryana. Indian journal of pathology & microbiology. 2010;53(2):308–9.

32. Ashraf H, Alam NH, Rothermundt C, Brooks A, Bardhan P, Hossain L, et al. Prevalence and risk factors of hepatitis B and C virus infections in an impoverished urban community in Dhaka, Bangladesh. BMC Infect Dis. 2010;10:208.

33. Athira KP, Vanathy K, Kulkarni R, Dhodapkar R. The prevalence of occult hepatitis B infection among the blood donors in a tertiary care hospital, Puducherry. Indian journal of medical microbiology. 2018;36(3):426–8.

34. Aziz S, Khanani R, Noorulain W, Rajper J. Frequency of hepatitis B and C in rural and periurban Sindh. JPMA The Journal of the Pakistan Medical Association. 2010;60(10):853–7.

35. Bhate P, Saraf N, Parikh P, Ingle M, Phadke A, Sawant P. CROSS SECTIONAL STUDY OF PREVALENCE AND RISK FACTORS OF HEPATITIS B AND HEPATITIS C INFECTION IN A RURAL VILLAGE OF INDIA. Arquivos de gastroenterologia. 2015;52(4):321–4.

36. Bhattar S, Aggarwal P, Sahani SK, Bhalla P. Co-Infections and Sero-Prevalence of HIV, Syphilis, Hepatitis B and C Infections in Sexually Transmitted Infections Clinic Attendees of Tertiary Care Hospital in North India. Journal of research in health sciences. 2016;16(3):162–5.

37. Chakravarti A, Roy P, Ashraf A, Siddiqui O. Trends in the epidemiology of hepatitis B virus infection at a tertiary care hospital: A 10-year retrospective analysis. Indian journal of gastroenterology : official journal of the Indian Society of Gastroenterology. 2015;34(2):189–90.

38. Dwibedi B, Sabat J, Ho LM, Singh SP, Sahu P, Arora R, et al. Molecular epidemiology of hepatitis B virus in primitive tribes of Odisha, eastern India. Pathogens and global health. 2014;108(8):362–8.

39. Faiz Ur R, Khan J, Fida Z, Parvez A, Rafiq A, Syed S. Identifiable risk factors in hepatitis B and C. J Ayub Med Coll Abbottabad. 2011;23(4):22–3.

40. Hafeez ud d, Siddiqui TS, Lahrasab W, Sharif MA. Prevalence of hepatitis B and C in healthy adult males of paramilitary personnel in Punjab. J Ayub Med Coll Abbottabad. 2012;24(3-4):138–40.

41. Jobayer M, Chowdhury SS, Shamsuzzaman SM, Islam MS. Prevalence of Hepatitis B Virus, Hepatitis C Virus, and HIV in Overseas Job Seekers of Bangladesh with the Possible Routes of Transmission. Mymensingh medical journal : MMJ. 2016;25(3):530–5.

42. Kazi AM, Shah SA, Jenkins CA, Shepherd BE, Vermund SH. Risk factors and prevalence of tuberculosis, human immunodeficiency virus, syphilis, hepatitis B virus, and hepatitis C virus among prisoners in Pakistan. International journal of infectious diseases : IJID : official publication of the International Society for Infectious Diseases. 2010;14 Suppl 3:e60–e6.

43. Khan F, Shams S, Qureshi ID, Israr M, Khan H, Sarwar MT, et al. Hepatitis B virus infection among different sex and age groups in Pakistani Punjab. Virology journal. 2011;8:225.

44. Khan MA, Zargar SA, Upadhyay J, Lone TA, Aggarwal R, Bashir G, et al. Epidemiology of hepatitis B and C viral infections in Ladakh region. Indian journal of gastroenterology : official journal of the Indian Society of Gastroenterology. 2018;37(6):504–10.

45. Khan S, Madan M, Virmani SK. Prevalence of Hepatitis B Virus, Genotypes, and Mutants in HBsAg-Positive Patients in Meerut, India. Iranian biomedical journal. 2019;23(5):354–61.

46. Khanani MR, Somani M, Khan S, Naseeb S, Ali SH. Prevalence of single, double, and triple infections of HIV, HCV and HBV among the MSM community in Pakistan. The Journal of infection. 2010;61(6):507–9.

47. Kinkel H-T, Karmacharya D, Shakya J, Manandhar S, Panthi S, Karmacharya P, et al. Prevalence of HIV, Hepatitis B and C Infections and an Assessment of HCV-Genotypes and Two IL28B SNPs among People Who Inject Drugs in Three Regions of Nepal. PLoS One. 2015;10(8):e0134455–e.

48. Kumari K, Seetlani NK, Akhter R. THE EMERGENT CONCERN OF SEROPOSITIVE STATUS OF HEPATITIS-B VIRUS AND HEPATITIS-C VIRUS IN THE PREGNANT FEMALES ATTENDING A TERTIARY CARE HOSPITAL. Journal of Ayub Medical College, Abbottabad : JAMC. 2015;27(1):155–7.

49. Makroo RN, Hegde V, Chowdhry M, Bhatia A, Rosamma NL. Seroprevalence of infectious markers & their trends in blood donors in a hospital based blood bank in north India. The Indian journal of medical research. 2015;142(3):317–22.

50. Meena M, Jindal T, Hazarika A. Prevalence of hepatitis B virus and hepatitis C virus among blood donors at a tertiary care hospital in India: a five-year study. Transfusion. 2011;51(1):198–202.

51. Mehta KD, Antala S, Mistry M, Goswami Y. Seropositivity of hepatitis B, hepatitis C, syphilis, and HIV in antenatal women in India. Journal of infection in developing countries. 2013;7(11):832–7.

52. Mishra KK, Trivedi A, Sosa S, Patel K, Ghosh K. NAT positivity in seronegative voluntary blood donors from western India. Transfusion and apheresis science : official journal of the World Apheresis Association : official journal of the European Society for Haemapheresis. 2017;56(2):175–8.

53. Pande C, Sarin SK, Patra S, Bhutia K, Mishra SK, Pahuja S, et al. Prevalence, risk factors and virological profile of chronic hepatitis B virus infection in pregnant women in India. Journal of medical virology. 2011;83(6):962–7.

54. Panigrahi R, Biswas A, Datta S, Banerjee A, Chandra PK, Mahapatra PK, et al. Anti-hepatitis B core antigen testing with detection and characterization of occult hepatitis B virus by an in-house nucleic acid testing among blood donors in Behrampur, Ganjam, Orissa in southeastern India: implications for transfusion. Virology journal. 2010;7:204.

55. Qureshi H, Bile KM, Jooma R, Alam SE, Afridi HUR. Prevalence of hepatitis B and C viral infections in Pakistan: findings of a national survey appealing for effective prevention and control measures. Eastern Mediterranean health journal = La revue de sante de la Mediterranee orientale = al-Majallah al-sihhiyah li-sharq al-mutawassit. 2010;16 Suppl:S15–S23.

56. Rauf A, Nadeem MS, Ali A, Iqbal M, Mustafa M, Latif MM, et al. Prevalence of hepatitis B and C in internally displaced persons of war against terrorism in Swat, Pakistan. European journal of public health. 2011;21(5):638–42.

57. Ray Saraswati L, Sarna A, Sebastian MP, Sharma V, Madan I, Thior I, et al. HIV, Hepatitis B and C among people who inject drugs: high prevalence of HIV and Hepatitis C RNA positive infections observed in Delhi, India. BMC public health. 2015;15:726.

58. Rudra S, Chakrabarty P, Hossain MA, Akhter H, Bhuiyan MR. Seroprevalence of Hepatitis B, Hepatitis C, HIV Infections in Blood Donors of Khulna, Bangladesh. Mymensingh medical journal : MMJ. 2010;19(4):515–9.

59. Rudra S, Chakrabarty P, Poddar B. Prevalence of hepatitis B and hepatitis C virus infection in human of Mymensingh, Bangladesh. Mymensingh medical journal : MMJ. 2011;20(2):183–6.

60. Shaha M, Hoque SA, Ahmed MF, Rahman SR. Effects of Risk Factors on Anti-HBs Development in Hepatitis B Vaccinated and Nonvaccinated Populations. Viral immunology. 2015;28(4):217–21.

61. Shamsuzzaman M, Singhasivanon P, Kaewkungwal J, Lawpoolsri S, Tangkijvanich P, Gibbons RV, et al. Hepatitis B among pregnant women attending health care facilities in rural Bangladesh. The Southeast Asian journal of tropical medicine and public health. 2011;42(6):1410–3.

62. Shanmugam RP, Balakrishnan S, Varadhan H, Shanmugam V. Prevalence of hepatitis B and hepatitis C infection from a population-based study in Southern India. European journal of gastroenterology & hepatology. 2018;30(11):1344–51.

63. Shedain PR, Baral G, Sharma KR, Dhital S, Devkota MD. Prevalence and Mother-to-newborn Transmission of Hepatitis B Virus in Tertiary Care Hospital in Nepal. Journal of Nepal Health Research Council. 2019;17(3):278–84.

64. Sheikh NS, Sheikh AS, Sheikh AA, Yahya S, Lateef M. Sero-prevalence of hepatitis B virus infection in Balochistan Province of Pakistan. Saudi journal of gastroenterology : official journal of the Saudi Gastroenterology Association. 2011;17(3):180–4.

65. Waheed A, Zafar ul A, Zaeem FA, Shariff MM, Qayyum A. Operating in a yellow nation; the frequency of hepatitis B and hepatitis C positive at a tertiary care teaching hospital. JPMA The Journal of the Pakistan Medical Association. 2010;60(12):1058–60.

